# Visual outcome of Anti-Vascular Endothelial Growth Factor Injections at the University College Hospital, Ibadan

**DOI:** 10.1101/2020.06.16.20132662

**Authors:** Tunji Sunday Oluleye, Yewande Olubunmi Babalola, Oluwole Majekodunmi, Modupe Ijaduola, Adeyemi Timothy Adewole

## Abstract

**Aim:** To evaluate the four-year outcome of intravitreal anti-vascular endothelial growth factor (anti-VEGF) therapy in an eye unit in sub-Saharan Africa.

**Methodology:** This retrospective study included 182 eyes of 172 patients managed in the vitreoretinal unit between 2016 and 2019 who were treated with intravitreal anti-VEGF Bevacizumab (1.25 mg/0.05 ml) with at least one year of follow up. The outcome measures were change in best-corrected visual acuity (BCVA) over one year of follow-up, the number of injections taken and complications.

**Results:** The mean age was 61.1 ± 16.3 years (M: F of 1:1.1) and about 62.1% were ≥ 60 years. A total of 330 injections were given during the period audited. The mean number of injections was 1.8 ± 0.93. Ninety-four (51.7%) eyes had only one injection while 33 (18.1%), 50 (27.5%) and 5 (2.7%) had 2, 3 and 4 injections, respectively. About 78.5% had moderate-severe visual impairment at baseline and 44.5%, 16.4%,12.6% and 7.1% at 1, 3, 6- and 12-months post injections, respectively. The mean BCVA improved for all eyes from 1.67 ±0.91 logMAR at baseline to 1.50±1.27 logMAR at one year. The logMAR letters gained was 23 at 1 month and 8.25 at 1 year with a statistically significant association between increasing number of injections and improved visual outcome (p= 0.015). One patient each developed endophthalmitis (0.6%) and inferior retinal detachment (0.6%) post-injection.

**Conclusion:** Visual acuity gain was recorded in patients who had at least two intravitreal Anti-VEGF injections in 1 year.

## Introduction

Vascular endothelial growth factor (VEGF) is stimulated in response to hypoxia in all vascularized intraocular tissue. The pathophysiologic mechanisms of most retinal diseases involve vascular proliferation and leakage in which VEGF is the main signal.^1^ Despite the complex pathophysiologic mechanisms of retinal damage in various intraocular vascular diseases, the use of anti-VEGF therapy is an effective therapeutic agent in various clinical trials for choroidal neovascular membrane from age-related macular degeneration (wet AMD), diabetic macular oedema (DME), macular oedema due to retinal vein occlusion (RVO), myopic choroidal neovascularisation (myopic CNV) and other retinal diseases.^2^

Anti-VEGF is also commonly used as standard therapy for various retinal vascular diseases, either as first or second-line treatment of choice.^2,3,4,5^ Although the use of Bevacizumab (Avastin) is off-label, it has been recommended as a cheaper non-inferior option to the other alternatives like Ranibizumab or Aflibercept. Similarly, bevacizumab is a preferred choice in low resource settings like ours, where patients in urgent need of anti-VEGF agents have to pay out of pocket.^6^

Various anti-VEGF treatment protocols have been advocated for the management of various retinal vascular diseases, with some dosing regimen supporting multiple injections.^4,5,7^ For instance, the “continuous dosing” regimen proposed by the MARINA study involves the use of monthly injections, while the PRONTO study offered “treatment as required” as guided by the OCT scan thus making patients require fewer injections unlike the MARINA study; but with similar visual outcome.^8,9^ Furthermore, the “treat and extend” regime aimed to reduce the treatment burden on patients by extending follow-up and/or treatment as determined by disease stability.^2,10^ The use of “initial 3 loading doses” followed by “as-needed treatment” showed excellent visual outcome in the ABC trial.^11^ Despite the cost-effectiveness of bevacizumab and proven treatment protocols recommending it to patients, the outcomes of the therapy have varied in the face of pervasive under treatment in low resource settings.

In this retrospective study, we seek to evaluate the 12-month visual outcome and relationship with the number of anti-VEGF injections in eyes with retinal vascular diseases to make recommendations for patients’ care in our practice and other similar settings to our health system where the scourge of retinal diseases is progressively becoming burdensome.^12,13^

## Methodology

In this retrospective study, medical records of patients who were treated with intravitreal bevacizumab at the University College Hospital, Ibadan, Nigeria from January 2016 to December 2019 were retrieved. This study adhered to the tenets of the Declaration of Helsinki. All patients consented to the Off-label use of bevacizumab. All procedures were carried out in the operating theatre under aseptic condition. The eye to be injected was prepared with 10% povidone-iodine to the periocular skin and lids, 5% povidone-iodine drops instilled into the conjunctiva sac, topical tetracaine was added before the povidone. Calliper measurements from the limbus were done to ensure injection was given through the pars plana (4mm for phakics and 3.5mm for pseudophakics; 1.5mm for neonates). Intravitreal injections (1.25 mg/0.05ml for adults, 0.625mg for neonates) of bevacizumab (Avastin, Roche, Basel, Switzerland) or 0.5 mg/0.05ml for adults, 0.25mg for neonates for ranibizumab (Lucentis, Novartis, Basel, Switzerland) were used. The choice of using either bevacizumab or ranibizumab was based on affordability by the patient. All patients were given topical moxifloxacin qid for 1 week after each injection.

Also, all patients were scheduled to have treatment on as-needed basis after the initial monthly 3 loading dosage. Eligible patients were those who had at least one injection. Patients were reviewed first-day post-injection for signs of undue inflammation and raised intraocular pressure. Patients were then reviewed monthly. Visual acuity assessment and dilated fundus examination were done at every visit. Optical Coherence Tomography scan was requested when visual acuity deteriorated.

Data extracted from case notes include clinical characteristics of the patients such as age, sex, eye (s) injected, systemic comorbidities, number of injections and best-corrected visual acuity (BCVA). The outcome measures were the change in BCVA over one year of follow-up, the number of injections taken and any associated complication.

Data was analysed using the Statistical Package for Social Sciences (SPSS), version 24 (SPSS Inc., Chicago, Illinois, USA), and reported as frequency distributions, percentages, and means ± standard deviation (SD). Snellen visual acuities were converted to logarithm of minimum angle of resolution (logMAR) for statistical analysis. BCVAs from the baseline, 1, 3, 6 and at 12 months were compared. Categorical variables were compared using χ^2^, and continuous variables were compared using the paired sample t-test. A p<0.05 was considered statistically significant.

## Results

A total of 206 patients’ medical records were requested but only 172 (83.5%) were retrieved successfully. One hundred and eighty-two eyes of 172 patients who had injections were analysed with a follow-up of 12 months. The mean age was 61.1 ±16.3 years (89 males, M: F of 1:1.1) with about 62.1% being 60 years and above (table 1). The most common indication was retinal venous occlusion, 64 (35.2%) followed by wet age-related macular degeneration, 42 (23%).

**Table 1:** Demographic and clinical characteristics of patients

|  | Frequency(n=182) | Percentage (%) |
| --- | --- | --- |
| **Age, mean =61.1± 16.3** |  |  |
| ≥ 60 years | 113 | 62.1 |
| 17-39 years | 6 | 3.3 |
| 40-59 years | 57 | 31.3 |
| 6-16 years | 2 | 1.1 |
| Infant-5years | 4 | 2.2 |
| **Sex** |  |  |
| Male | 89 | 48.9 |
| Female | 93 | 51.1 |
| **Eye** |  |  |
| Right Eye | 94 | 51.6 |
| Left Eye | 88 | 48.4 |
| **Indications** |  |  |
| Wet Age-related macular degeneration/CNVM | 42 | 23.0 |
| CRVO | 34 | 18.7 |
| PDR/DME | 27** | 14.8 |
| IPCV | 18 | 9.9 |
| HRVO | 16 | 8.9 |
| BRVO | 14 | 7.7 |
| CME | 9 | 4.9 |
| SCR | 6 | 3.3 |
| RAM | 4 | 2.2 |
| ROP | 3 | 1.7 |
| NVG | 2 | 1.0 |
| Others | 7*** | 3.9 |
| **Baseline Snellen Visual Acuity (logMAR)** |  |  |
| 6/6 (0.0) | 5 | 2.7 |
| 6/9 (0.2) | 4 | 2.2 |
| 6/12 (0.3) | 7 | 3.8 |
| 6/18 (0.5) | 16 | 8.8 |
| 6/24 (0.6) | 8 | 4.4 |
| 6/36 (0.8) | 17 | 9.3 |
| 6/60 (1.0) | 16 | 8.8 |
| CF (2.3) | 77 | 42.3 |
| HM (2.5) | 25 | 13.7 |
| LP (3.0) | 2 | 1.1 |
| NLP (4.0) | 2 | 1.1 |

**Table 1: Demographic and clinical characteristics - continued**
|  | Frequency(n=182) | Percentage (%) |
| --- | --- | --- |
| **No of injections**, mean=1.8 ± 0.93 |  |  |
| 1 | 94 | 51.7 |
| 2 | 33 | 18.1 |
| 3 | 50 | 27.5 |
| 4 | 5 | 2.7 |
| **Type of Injection** |  |  |
| Bevacizumab (Avastin) | 180 | 98.9 |
| Ranibizumab (Lucentis) | 2 | 1.1 |
| **Complications** |  |  |
| Endophthalmitis | 1 | 0.6 |
| Retinal detachment * | 1 | 0.6 |

CNVM= choroidal neovascular membrane; AMD= age related macular degeneration; CRVO= central retinal vein occlusion; PDR= proliferative diabetic maculopathy; DME= diabetic macular oedema; IPCV= Idiopathic Polypoidal Choroidal Vasculopathy; HRVO= hemi-retinal vein occlusion; BRVO= branch Retinal Vein Occlusion; CME= cystoid macular oedema; SCR= sickle cell Retinopathy; RAM= retinal arterial macroaneurysm; ROP= retinopathy of prematurity; NVG= neovascular glaucoma
\* 3 infants with Retinopathy of Prematurity had no objective assessment of visual acuity of documented
\*\* one patient had severe non-proliferative diabetic retinopathy
\*\*\*Vitreous haemorrhage in POAG, Presumed Toxoplasmosis, Choroidal melanoma, Intraretinal mass (unspecified), Haemorrhagic macular detachment, Specific uveitis and Myopic CNVM
\* Inferior bullous developed post 3rd injection

About 78.5% had moderate-severe visual impairment (MSVI) at baseline and 44.5%, 16.4%,12.6% and 7.1% at **1**, 3, 6 and 12months post injections respectively (Figure 1). A total of 330 injections were given with the mean number of injections of 1.8 ± 0.93. Ninety-four eyes, 94 (51.7%) eyes had only one injection while 88 (48.3%) had at least 2 injections.

**Figure 1:**
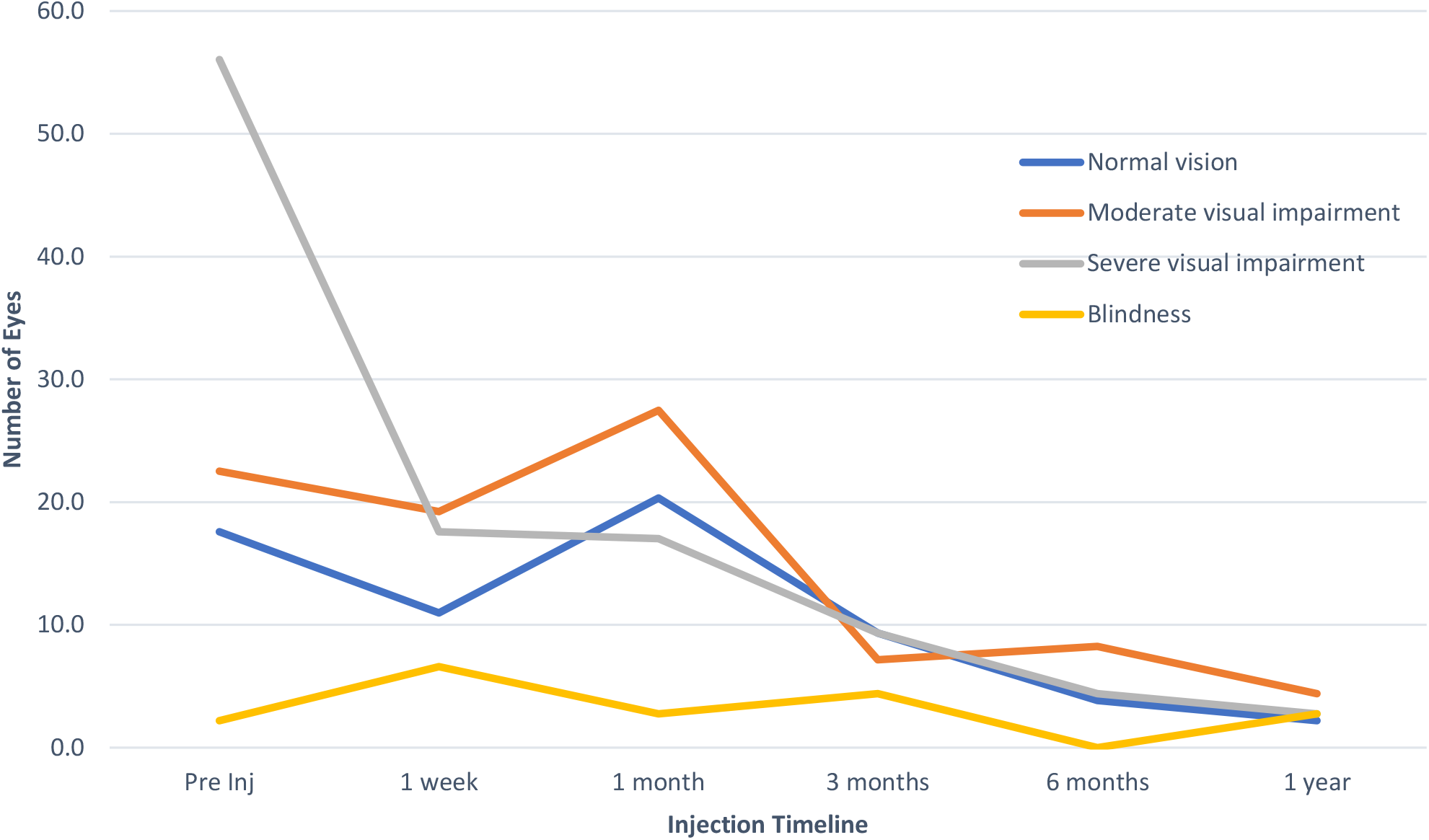
Trend in visual function with follow-up.

The mean BCVA improved for all eyes from 1.67 ±0.91 logMAR at baseline to 1.21 ±1.01 logMAR, 1.41 ± 1.09 logMAR, 1.44 ±1.17 logMAR and 1.50±1.27 logMAR at 1 month, 3 months, 6 months and one year, respectively. The logMAR letters gained progressively declined from 23 at 1 month to about 8 letters at 1 year (Figure 2). Patients who had at least 2 injections were significantly (p=0.015) more likely to have better visual outcome than those who had just one injection (table2). One patient each developed endophthalmitis (0.6%) and inferior retinal detachment (0.6%) post-injection.

**Figure 2:**
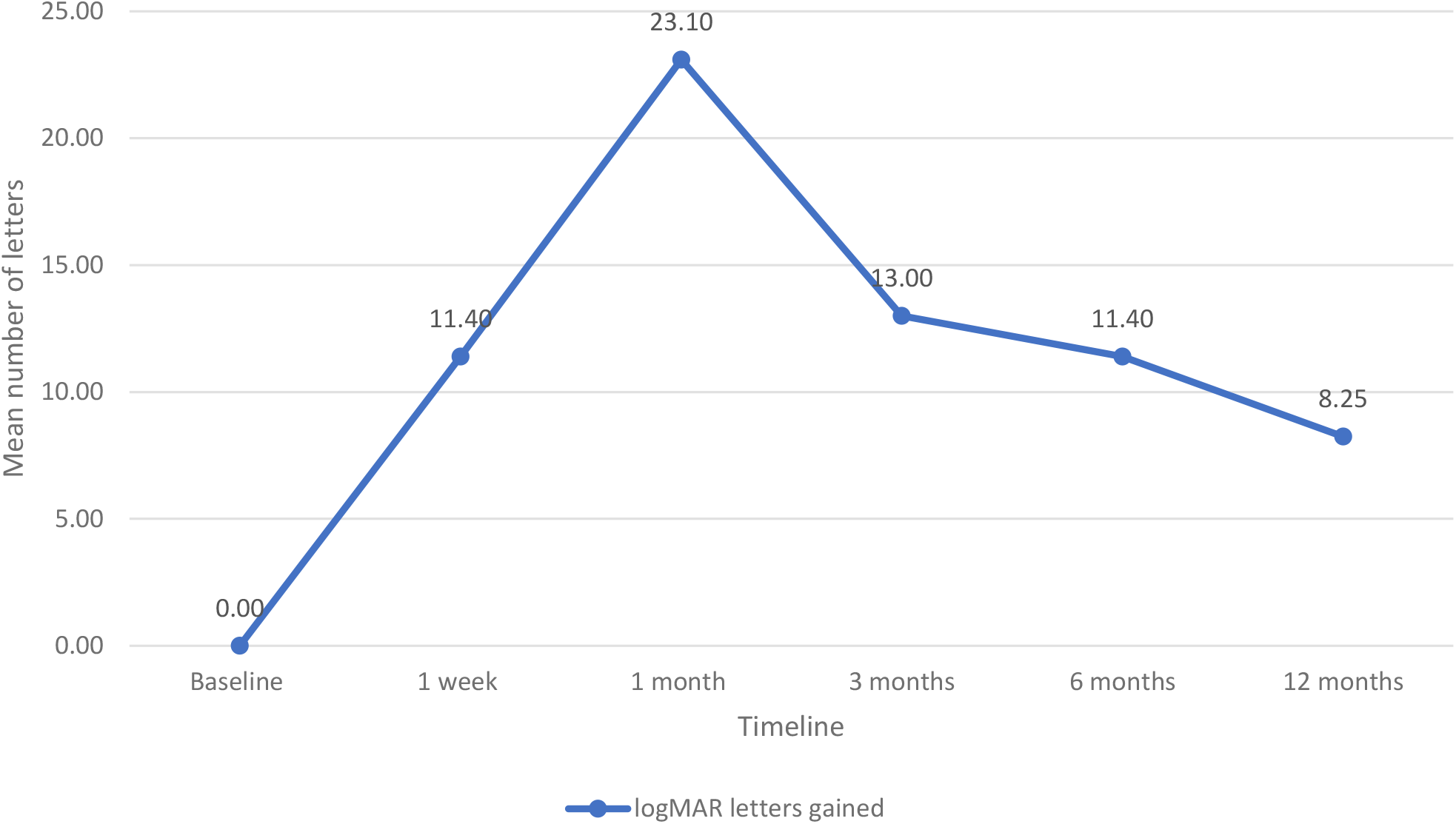
Trend analysis of mean change in mean best corrected visual acuity represented by letters gained.

**Table 2:** Relationship between clinical characteristics and number of injections

|  | One Injection | Two to four Injections | X<sup>2</sup> | p-value |
| --- | --- | --- | --- | --- |
|  | N (%) | N (%) |  |  |
| **I week post-injection** |  |  |  |  |
| Better than 0.8 log MAR (6/6 to ≥ 6/60) | 31 (35.6) | 56 (64.4) | 17.121 | **0.039\*** |
| 0.8 logMAR or worse (< 6/60) | 63 (66.3) | 32 (33.7) |  |  |
| **1-month post-injection** |  |  |  |  |
| Better than 0.8 logMAR (6/6 to ≥ 6/60) | 31 (35.6) | 56 (64.4) | 17.121 | **0.039\*** |
| 0.8 logMAR or worse (< 6/60) | 63 (66.3) | 32 (33.7) |  |  |
| **3 months post-injection** |  |  |  |  |
| Better than 0.8 logMAR (6/6 to ≥ 6/60) | 7 (23.3) | 23 (76.7) | 11.532 | **0.001\*** |
| 0.8 logMAR or worse (< 6/60) | 87 (57.2) | 65 (42.8) |  |  |
| **6 months post-injection** |  |  |  |  |
| Better than 0.8 logMAR (6/6 to ≥ 6/60) | 6 (27.3) | 16 (72.7) | 5.754 | **0.015\*** |
| 0.8 logMAR or worse (< 6/60) | 88 (55.0) | 72 (45.0) |  |  |
| **12 months post-injection** |  |  |  |  |
| Better than 0.8 logMAR (6/6 to ≥ 6/60) | 2 (16.7) | 10 (83.3) | 6.294 | **0.015\*** |
| 0.8 logMAR or worse (< 6/60) | 92 (54.1) | 78 (45.9) |  |  |
| **Sex** |  |  |  |  |
| Female | 44 (47.3) | 49 (52.7) | 1.432 | 0.231 |
| Male | 50 (56.2) | 39 (43.8) |  |  |
| **Eye** |  |  |  |  |
| Left | 46 (51.7) | 43 (48.3) | 0.000 | 0.992 |
| Right | 48 (51.6) | 45 (48.4) |  |  |
| **Age** |  |  |  |  |
| ≥ 60 years | 53 (46.9) | 60 (53.1) | 7.103 | 0.115 |
| ≥ 60 years | 41 (53.1) | 28 (46.9) | | |

- \* Statistically significant level (p < 0.005)

## Discussion

The mean age of patients involved in this study is similar to that in other centres in Nigeria.^14–17^ But, the wide age range was due to the inclusion of babies who received anti-VEGF for retinopathy of prematurity.

Almost all our patients,180 (98.9%) had the less expensive bevacizumab injection, which is akin to other studies done both locally.^14–17^ On the other hand, a report from the oil-rich setting of Port-Harcourt in Nigeria reported about 25% usage of Ranibizumab.^18^ A vial of bevacizumab is shared (repackaged bevacizumab) by 10-20 patients to further reduce the cost in a pooled funding system of the unit to encourage compliance with the treatment regime. Even though our patients had been counselled on the need to have the recommended monthly dose of 3 injections, only about 27.5% (50) complied to regimen and 51.7% (94) had only one injection due to financial constraints as they pay out of pocket. The average number of injections in this cohort (1.8 ± 0.9) is akin to a similar study reported in a tertiary hospital in Benin,^16^ but at variance with the higher mean number of injections reported in major landmark clinical trials that have shaped the preferred practice pattern for intravitreal anti-VEGF injections.^2,8–10,12^ A strong health system with an effective financial payment and insurance coverage could have been the reason for this disparity.

We found that this cohort had improved vision which was better in those that had at least two injections over the 12 months follow-up period. The average number of letters gained was highest at one-month post-injection but was not sustained as it declined to 13 and finally to 8.5 letters (1.7 lines) at 12-month but this improvement was still a statistically significant outcome. Uhumwangho^16^ similarly reported visual improvement in about 50% of studied patients. Pham et al,^2^ in their systematic review and meta-analysis reported varying levels of visual outcomes depending on the specific disease. Chin-Yee et al,^4^ systematic reviews reported a mean improvement in visual acuity of 5.4 and about 10 ETDRS letters in PRN and treat and extend regimens respectively and the average of 5.6 and 8 injections over 12 months for wet ARMD. Lai et al^20^ reported 14 letters (2.8 lines) improvement for myopic CNVM at 24 months. Conversely, Low et al,^7^ stated that among the 17 trials reviewed, none of them stated any clinically significant visual improvement (> 5 letters) across the three major retinal vascular diseases scheduled for anti-VEGF injections (wet ARMD, DME and Central/Branch retinal vein occlusion irrespective of the type of anti-VEGF agents used (bevacizumab, ranibizumab and aflibercept).

## Conclusion

Our review has shown that the visual gain was more in those who had at least 2 injections. Therefore, it is recommended that anti-VEGF agents should be made available on the National Health Insurance Scheme, as well as enrolment of all citizens on the scheme. These acts will among other things, make anti-VEGF available and accessible to those that need it, thereby aiding to ease the various burdens associated in accessing these drugs.

## Limitations

The study being a retrospective study, may be limited by accurate data retrieval from case records.

## Data Availability

Data from patients' medical records were inputted into SPSS for analysis.

## Contributions

I thank Mr. Iwa of the records department for the excellent work done in retrieving the case notes. Appreciation goes to Dr. Ibiyemi (Senior Registrar, Ophthalmology) and Mr. Seun Ayangbesan (Research assistant) who helped in data extraction. TO, YOB, OM, MI, and AA contributed to the design of the study and. AA analysed the data and prepared the initial manuscript. All authors reviewed, proofread, and approved the final manuscript and references.

